# Testing real-world feasibility and acceptability of implementing an intravenous iron intervention for pregnant women with moderate and severe anaemia in Bangladesh: A demonstration project protocol

**DOI:** 10.1101/2025.01.22.25320952

**Authors:** Khic-Houy Prang, A.M Quaiyum Rahman, Ebony Verbunt, Hana Sabanovic, Shamim Ahmed, Mohammed Imrul Hasan, Eliza M. Davidson, Alistair R.D McLean, Sabine Braat, Clare Glover-Wright, Natalie Carvalho, Sant-Rayn Pasricha, Jena D. Hamadani, Bidhan K. Sarker

**Affiliations:** Centre for Health Policy, Melbourne School of Population and Global Health, The University of Melbourne, Melbourne, Australia; Maternal and Child Health Division, International Centre for Diarrhoeal Disease Research, Bangladesh (icddr,b), Dhaka, Bangladesh; Population Health and Immunity Division, Walter and Eliza Hall Institute of Medical Research, Melbourne, Australia; Centre for Epidemiology and Biostatistics, Melbourne School of Population and Global Health, The University of Melbourne, Melbourne, Australia

**Keywords:** anaemia, intravenous iron, maternal health, demonstration project, implementation research, Bangladesh, low- and middle-income countries

## Abstract

**Background:** Antenatal anaemia is a significant global health problem, affecting 48% of pregnant women in South Asia. The standard of care in Bangladesh is oral iron and folic acid supplementation. However, access and adherence to oral iron supplementation are subpar. An alternative treatment available to address antenatal anaemia is intravenous (IV) iron. Modern IV iron products are routinely used in high-income clinical settings, including primary and secondary care, to deliver a high dose of iron in a single short infusion. A demonstration project will be conducted to test the real-world feasibility and acceptability of implementing an IV iron intervention to treat pregnant women with moderate and severe anaemia in the primary healthcare setting of Bangladesh. In this protocol paper, we describe the implementation research program that will guide the development, implementation, and evaluation of an IV iron intervention in real-world settings.

**Methods:** We will use implementation science frameworks to guide the demonstration project. The implementation research program includes three phases: 1) a formative phase in preparation for the implementation of an IV iron intervention involving a review, qualitative research, and readiness assessment; 2) the development and implementation of IV iron intervention care pathways and strategies to support the uptake and delivery of the intervention using a participatory research approach; and 3) a process evaluation of IV iron intervention care pathways and strategies involving qualitative and quantitative assessment of the costs, processes and contextual factors affecting its implementation.

**Discussion:** Modern IV iron products present a novel opportunity to reduce the disproportionate burden of antenatal anaemia in low-and-middle income countries (LMICs). The demonstration project will ascertain whether an IV iron intervention can be effectively introduced into routine antenatal care in Bangladesh. The extent to which it is considered an acceptable treatment by pregnant women with moderate or severe anaemia receiving the intervention, healthcare providers delivering the intervention, and policymakers will be determined. If successful, understanding how an IV iron intervention will be implemented across several care pathways and its associated costs will inform the scalability of an IV iron intervention in the primary healthcare system of Bangladesh and provide implementation guidance in other LMICs.

**Contributions to the literature:**

- Modern IV iron products can quickly restore iron levels, offering a new opportunity to address antenatal anaemia in LMICs.
- The demonstration project in Bangladesh will involve pregnant women, local communities, and healthcare workers to co-design anaemia care pathways and strategies for implementing an IV iron program in primary care.
- The demonstration project provides access to an alternative treatment, trains healthcare workers, and builds local capacity for anaemia management.
- Testing and refining anaemia care pathways and strategies in a real-world setting will offer insights for scaling and adapting these approaches to other regions or population, creating a template for similar initiatives.

## Background

Antenatal anaemia is a significant global health problem with the highest burden disproportionately affecting women in South Asia (48%), West and Central Africa (52%) (1). The World Health Organisation (WHO) defines antenatal anaemia as a haemoglobin level of less than 11 g/dL (2). The most common cause of anaemia is iron deficiency (3), accounting for approximately half of all antenatal anaemia cases worldwide (4, 5). Iron deficiency anaemia is associated with significant adverse health outcomes including pre-eclampsia, haemorrhage, premature birth, low birth weight infants, and maternal mortality (5, 6). In 2012, the WHO set an ambitious global nutrition target of halving anaemia prevalence in women of reproductive age by 2025 (7), and later extending the target to 2030 due to limited progress (8).

In low and middle-income countries (LMICs), oral iron and folic acid (IFA) supplementation is a common low-cost intervention to prevent and treat iron deficiency anaemia among pregnant women. WHO recommends daily IFA supplementation throughout pregnancy (9). IFA supplementation is generally efficacious, though coverage and adherence are suboptimal (10). In Bangladesh, IFA has been part of routine antenatal care and included in the essential medicine list since 2001 (11), with 74% of pregnant women consuming IFA tablets. However, only 46% consumed the recommended intake of ≥90 IFA tablets (12) due to factors such as limited access and supply of IFA, gastrointestinal side-effects, late antenatal care, and misconceptions about IFA resulting in large babies and caesarean sections (13, 14). Subsequently, the prevalence of anaemia in pregnant women in Bangladesh remains high at 42% (15), with a higher prevalence rate in rural areas (44%) compared to urban areas (35%) (16).

Modern intravenous (IV) iron formulations such as ferric carboxymaltose (FCM) are safe and effective and are increasingly used in high income countries (HICs) as second-line treatment to treat iron deficiency anaemia in pregnant women from the second trimester onwards when there is poor response to comply with oral iron (17, 18). These formulations can be given rapidly in a single short infusion with anaphylaxis extremely rare (<1%) (18, 19). Most recently, several reviews have proposed the use of parenteral iron as first-line treatment for iron deficiency anaemia in pregnancy, particularly for second and third trimesters (20, 21).

Modern IV iron formulations provide an opportunity to reduce the disproportionate burden of antenatal anaemia in LMICs. However, the challenge lies in adapting the IV iron intervention from HICs to LMICs, where healthcare systems are resource constrained and sociocultural factors could hinder widespread implementation (22). Considering the aforementioned factors and that the translation of research evidence into public health policy and practice can take up to 17 years (23), a demonstration project will be conducted to ‘demonstrate’ whether IV iron can be implemented in the primary healthcare system of Bangladesh. Demonstration projects are often used to test the acceptability, feasibility, implementation, and scalability of evidence-based interventions in a specific context or population, and to gather data on the barriers and facilitators to implementation in real-world settings, where the intervention may be later introduced. The results of the demonstration project can inform the development of larger implementation studies and guide decision-making about the implementation of the intervention in other settings.

## Aims

The objective of this paper is to describe the protocol for a demonstration project which aims to test the real-world feasibility and acceptability of implementing an IV iron intervention to treat pregnant women with moderate and severe anaemia in the primary healthcare setting of Bangladesh. Specifically, we aim to:

1. understand how antenatal anaemia is currently managed in the study setting, and identify potential health system barriers and enablers to implementing an IV iron intervention;
2. co-design antenatal anaemia screening and referral pathways, and culturally appropriate strategies with pregnant women, community members, and healthcare providers to inform the implementation of an IV iron intervention in primary care; and
3. implement and evaluate the feasibility, acceptability and costs of antenatal anaemia screening and referral pathways and strategies to increase anaemia screening and IV iron treatment among pregnant women.

The demonstration project is part of a larger program of research “Efficacy and Demonstration of IntraVenous Iron for Anaemia in pregnancy” (EDIVA) which aims to assess the effectiveness and feasibility of IV iron (FCM) to treat pregnant women with moderate and severe anaemia in the second or third trimester in Bangladesh. This includes an antenatal anaemia prevalence survey (24, 25), a randomised controlled trial (RCT) to determine the efficacy and safety of IV iron compared to standard of care oral iron supplementation (registration number ACTRN12621000968875) (26), and this demonstration project. The RCT and demonstration project will be accompanied by a micro-costing study and economic evaluation to understand the costs associated with IV iron implementation, and the cost-effectiveness of IV iron compared to oral iron supplementation.

The proposed demonstration project involves a multi-phase implementation research program using different methods and approaches to address our previously stated aims: 1) a formative phase in preparation for the implementation of an IV iron intervention involving a review, qualitative research, and readiness assessment; 2) the development and implementation of IV iron intervention care pathways and strategies to support the uptake and delivery of the intervention using a participatory research approach; and 3) a process evaluation of IV iron intervention care pathways and strategies involving qualitative and quantitative assessment of the processes and contextual factors affecting its implementation (Figure 1).

**Figure 1.**
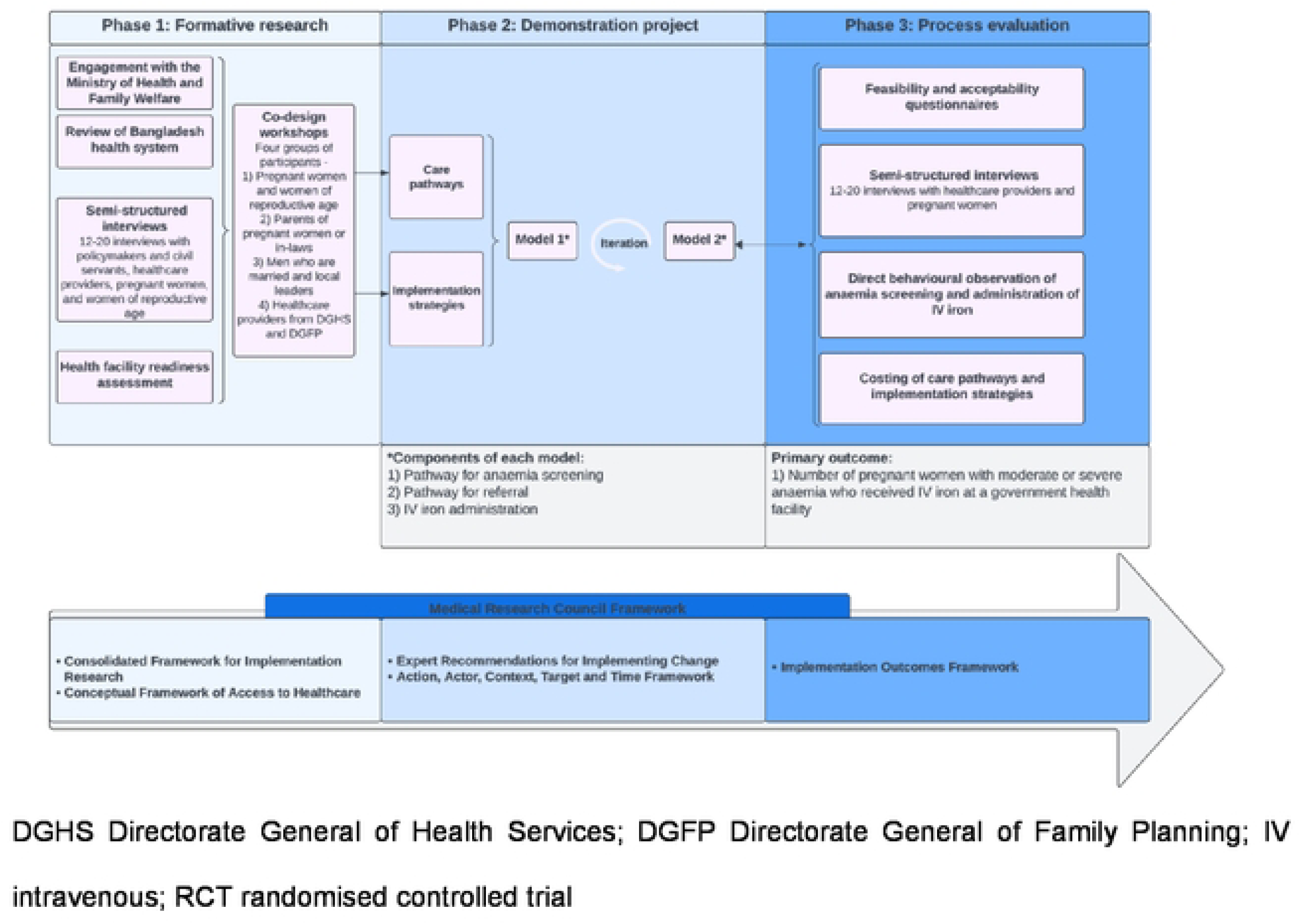
Demonstration project multi-phase implementation research program

## Methods

### Study setting

The study will be conducted in Bangladesh, a densely populated country with a total population of 171.2 million as of 2022 (27). In 2022, people aged 15-64 years accounted for 68% of the total population, with 65% of women of reproductive age (15-49 years) (27).

The health system comprises four key actors: public sector (government), for-profit private sector, not-for-profit private sector (non-governmental organisations), and international development organisations. The Ministry of Health and Family Welfare, through the Directorate General of Health Services (DGHS) and Directorate General of Family Planning (DGFP), is responsible for the delivery of general health and family planning services across three levels of healthcare linked by a referral system: 1) three tiers of primary care (Community Clinics at the ward level; Rural Health Centers, Union Sub-Centers, and Union Health and Family Welfare Centers at the union level; and Upazila Health Complexes at the upazila level); 2) secondary care (District Hospitals, Mother and Child Welfare Centres at the district level) and 3) tertiary care (specialised healthcare centres, medical college hospitals in large urban centers at the division level).

The demonstration project will be conducted in Bandar Upazila of the Narayanganj District in central Bangladesh. Bandar is divided into 172 villages across five unions: Bandar, Dhamgar, Kalagachhia, Madanpur, and Musapur. In 2023, the population was 236,190 with 50,011 (21%) women of reproductive age (15-49 years) and an estimated 2,368 (5%) of pregnant women per year (unpublished data, Family Planning Office Bandar Upazila). The prevalence of anaemia among pregnant women in Bandar Upazila is 37%, similar to adjacent Rupganj (40%) and Sonargoan (37%) Upazilas (24, 25). Maternal, newborn, and child health services are delivered across 14 Community Clinics, 1 Union Health Sub-Centre, 4 Union Health and Family Welfare Centres, 1 Upazila Health Complex, 1 Mother and Child Welfare Centre, and 1 District Hospital.

### Research program

In this study protocol, we outline the implementation research activities underpinned by the UK Medical Research Council (MRC) framework for developing and evaluating complex interventions (28, 29) and implementation science frameworks (e.g. Consolidated Framework for Implementation Research and Implementation Outcomes Framework) that will guide the development of a demonstration project. Using a post research design (30), a demonstration project will be conducted to assess multiple anaemia screening, referral and treatment pathways, and implementation strategies in the primary healthcare system of Bangladesh. Research will be carried out in three phases: 1) formative research, 2) demonstration project, and 3) process evaluation. During the formative research, we will investigate Bangladesh’s sociocultural environment and the capacity of its health system and policy pathways before the introduction of an IV iron intervention. The results will guide the development and implementation of the demonstration project, which will operate for one year. Costs to government, including providers, and to pregnant women and their families, will be collected alongside the demonstration project. The demonstration project will be followed by a mixed methods process evaluation. Detailed overviews of each phase are described in the following section using the Standards for Reporting Implementation Studies (STaRI) checklist (Appendix 1).

### Phase one: Formative research

#### Health system review

Guided by Berman and Bitran’s health system framework (31), we will conduct a health system review of Bangladesh to identify factors that influence how an IV iron intervention can be effectively integrated within routine antenatal care services in the primary healthcare setting. This will include a search of scientific databases and a grey literature search with documents review of local, state, and national maternal and child health policies programs and funding structures. Findings will be described according to the key dimensions of the health system: organisation structure, financing, regulation and planning, physical and human resources, provision of services, and principal healthcare reforms.

Exploring the Bangladesh health systems context will allow for a comprehensive understanding of the structure and performance of the health system and the roles played by government agencies, the private sector, and non-governmental organisations in delivering maternal and child health programs.

#### Key stakeholders’ interviews

We will conduct semi-structured interviews with key stakeholders to identify constraints and resources available and the most promising care pathways for incorporating an IV iron intervention in the primary healthcare system and the support required to do this. Key stakeholders will include: 1) policymakers and civil servants from DGHS, DGFP and Directorate General of Drugs Administration (DGDA); 2) healthcare providers; and 3) pregnant women and women of reproductive age (15-49 years).

##### Data collection and analysis

We will approach policymakers, civil servants and healthcare workers who currently operate within the public healthcare system of Bangladesh and have experience developing and implementing maternal and child health programs and invite them to be interviewed. We may use snowball sampling in which participants may be asked to identify and refer colleagues who would be a good fit for the research. Pregnant women and women of reproductive age who completed the anaemia prevalence survey (24, 25) will be identified and invited to participate in the interview.

Informed by CFIR and the Conceptual Framework of Access to Healthcare (32), we will develop separate interview guides for each of the participant groups focusing on contextual and process factors likely to influence the implementation of an IV iron intervention in routine antenatal care and opportunities for upscaling. All interview guides will be translated into Bengali (the national language of Bangladesh).

Past qualitative research has shown that saturation of themes is likely to be achieved with approximately 12–20 interviews in a homogenous group (33, 34). As such, we require a minimum of 12 participants for each qualitative study to ensure that there is sufficient data to allow for qualitative assessment of key themes. We will audio-record, transcribe interviews and cross-check all transcriptions to ensure validity. We will apply the framework method using a mix of deductive and inductive thematic analysis to identify themes (35). We will report the qualitative studies using the Standards for Reporting Qualitative Research reporting guideline (36).

#### Health facilities readiness assessment

We will conduct a readiness assessment of health facilities in Bandar Upazila to describe the characteristics of the health facilities including the detection and management of antenatal anaemia care processes. Assessing service specific readiness will provide information on available and functional equipment, additional resources required and the capacity of the government and healthcare workforce to effectively adopt and deliver an IV iron intervention within the primary healthcare system.

##### Data collection and analysis

We will collect cross-sectional data by using an interview-administered questionnaire adapted from the WHO Service Availability and Readiness Assessment Standard tool (SARA) (37) and the Bangladesh health facility assessment tool. The readiness assessment will take place in all 22 government health facilities in Bandar Upazila and will include nine modules: 1) general characteristics, 2) facility medical records and data management systems, 3) basic amenities, 4) infection control, 5) protocols/guidelines and training, 6) equipment – antenatal care, 7) equipment – IV iron, 8) emergency care – IV iron, and 9) supplementation. All questionnaire data will be verified by a structured facility observation checklist. Service specific readiness for antenatal anaemia care and IV iron administration will be determined by mean readiness index scores across several domains.

#### Co-design workshops

Using a co-design approach, we will develop care pathways and implementation strategies with end-users, community members, and healthcare providers to support the implementation of an IV iron intervention in primary care. Co-design aims to equitably involve all stakeholders (e.g., researchers, healthcare providers, end-users) in the design process to co-develop solutions to improve the quality and the delivery of healthcare services.

The co-design approach will be conducted in two stages: 1) an information gathering stage to understand the ‘touchpoints’ (i.e., any point of contact people has with a service) among end-users, community members, and healthcare providers and 2) co-design workshops where end-users jointly develop the care pathways and implementation strategies with healthcare providers and the research team. The formative research (i.e., health system review, semi-structured interviews with policymakers, pregnant women and healthcare providers, and health facilities service readiness) will constitute an information gathering stage that identifies the touchpoints, barriers and enablers, and further develops an in-depth understanding of these issues from multiple perspectives. In stage two, a summary of the data collected in stage one will be presented to participants in the co-design workshops to inform the subsequent discussion. End-users, healthcare providers, and the research team will then work together to co-develop solutions to the identified areas for improvement through a series of structured and facilitated workshops (38, 39).

##### Data collection and analysis

Co-design workshops will be held in Bandar Upazila with four groups of participants: 1) pregnant women and women of reproductive age (15-49 years old); 2) parents of pregnant women or in-laws; 3) men who are married (with or without children) and local leaders (formal leadership role in the community such as Imams); and 4) healthcare workers from DGHS and DGFP

Participants will be purposively sampled using maximum variation (cases that are different to each other) to capture as many different viewpoints as possible. Following the completion of each semi-structured interview, pregnant women and healthcare workers who appeared to be interested in providing more feedback will be invited to participate in the co-design workshops. Additional pregnant women will be recruited via the prevalence survey. Using snowballing technique, family members of the pregnant women including their husbands and mothers-in-law will be invited. We will identify local leaders through icddr,b networks, and local knowledge, and approach them directly. In each group, 15 participants will be invited to participate with the aim of recruiting 10 participants per workshop. Previous case studies and review have reported between 2 and 16 participants per co-design workshop (40, 41).

Co-design meetings will be facilitated by a member of the research team. The Consolidated Framework for Implementation Research–Expert Recommendations for Implementing Change matching tool (42) will inform the development of strategies. The selection of the care delivery pathways and implementation strategies will be based on stakeholder consensus and research team feedback and rated against the APEASE criteria: affordability; practicability; effectiveness and cost-effectiveness; acceptability; side-effects and safety; and equity (43). The action plan will be mapped to the Action, Actor, Context, Target, and Time (AACTT) framework (44) to specifically describe the behaviours of multiple actors across different levels of the organisation required to enact change.

### Phase two: Demonstration project

#### Anaemia screening

WHO recommends full blood count (FBC) testing as the preferred method for diagnosing anaemia during pregnancy (9), with serum ferritin concentrations an indicator for iron deficiency anaemia (45). However, FBC testing is not always readily available in primary care settings in LMICs. Given that there are currently no point-of-care assays that can determine the presence of iron deficiency in real time in the field, on-site Hb testing of capillary blood with a haemoglobinometer is recommended over the use of the Hb colour scales as the method for diagnosing anaemia in pregnancy (9). Using point-of-care testing will enhance the feasibility of delivering an IV iron intervention in primary care and enable the translation of the findings in low-resource settings.

Pregnant women will be screened for anaemia using point-of-care testing (i.e., HemoCue) during the second and third trimester of pregnancy (week 13 to week 32 of pregnancy) as identified by date of last menstrual period. All finger pricks and Hb assessments will be performed by government healthcare workers. After swabbing the skin using alcohol solution, a puncture will be made on the tip of the middle, ring, or index finger using a disposable lancet. A single drop of blood will be placed on the cuvette and inserted into the HemoCue device to determine the Hb level. Pregnant women found to be moderately or severely anaemic (Hb <10 g/dL) will be referred to attend the appropriate site for the delivery of IV iron within the next week.

#### IV iron intervention

FCM is approved by the DGDA for clinical use in Bangladesh. We will procure Vifor FCM from the manufacturer directly (CSL Vifor). Government healthcare workers will be trained and mentored by our research team to infuse IV iron to pregnant women with anaemia in the primary healthcare setting.

Following informed consent, eligible pregnant women will complete a questionnaire to collect basic demographic and medical information. A trained government health worker will perform a physical examination (as per standard antenatal care procedures), and then administer IV iron, with oversight from an icddr,b trained physician. Pregnant women will receive FCM – 20mg/kg up to 1000mg (women 50kg or above) in 250mL normal saline – intravenously over 15 minutes. An intravenous cannula will be inserted following a standard aseptic procedure. The skin will be cleaned with ethanol, and a sterile cannula will be inserted into the forearm or hand by a trained nurse, and the cannula will be fixed in place with a sterile Tegaderm or clinical tape. Pregnant women will be monitored over the 15 minutes of the infusion for any adverse events, and if they develop, these will be attended promptly and treated according to standard clinical management guidelines. Pregnant women will be observed for a further 45 minutes. Following completion, the cannula will be removed and placed in a biohazard container, and a band-aid applied to the arm. We will follow universal precautions whilst working with sharps. Administration of iron will be conducted in a room equipped with a ‘crash’ trolley which will contain adrenaline, hydrocortisone, intravenous fluids, and antihistamines and oxygen cylinders for emergencies. The contents and expiry of the crash trolley and availability of oxygen will be confirmed each day by healthcare workers before administration of the drug.

We will provide the sites with anaemia screening tools, FCM iron formulation, IV iron medical instruments, oxygen cylinder and a crash trolley if found to be not available following the health facilities’ readiness assessment. Where possible, we will use existing resources and infrastructures in the primary healthcare setting to reflect real-world routine conditions.

#### Care pathways

We developed two different anaemia screening, diagnosis, referral, treatment approaches for the demonstration project. The findings from phase 1 will be reported in a separate paper.

##### Study design

The demonstration project will be a post research design (30) in a new setting that assesses the feasibility and acceptability of several care delivery pathways co-designed by community members and healthcare workers, without any usual care comparison group. We will conduct a staggered roll-out of an IV iron intervention in Bandar Upazila to ensure sufficient resources and support are distributed to each site throughout the implementation of the demonstration project.

##### Study participants and recruitment

Pregnant women in the second or third trimester who attend a primary healthcare facility at the ward, union, or upazila level in Bandar Upazila will be invited to participate in the study by a healthcare worker. The healthcare worker will provide educational information about antenatal anaemia and the study. Healthcare workers (e.g., family welfare assistants and health assistants) who routinely conduct community outreach activities in catchment villages including visiting women of reproductive age, pregnant women, and eligible couples for family planning services will provide information about the study. We will also identify and recruit pregnant women using family welfare assistants’ register. The register contains information on individual pregnancy as women of reproductive age are screened regularly for pregnancy (via questioning on last menstrual period) by family welfare assistants. Participant inclusion and exclusion criteria are described in Table 1.

**Table 1.**
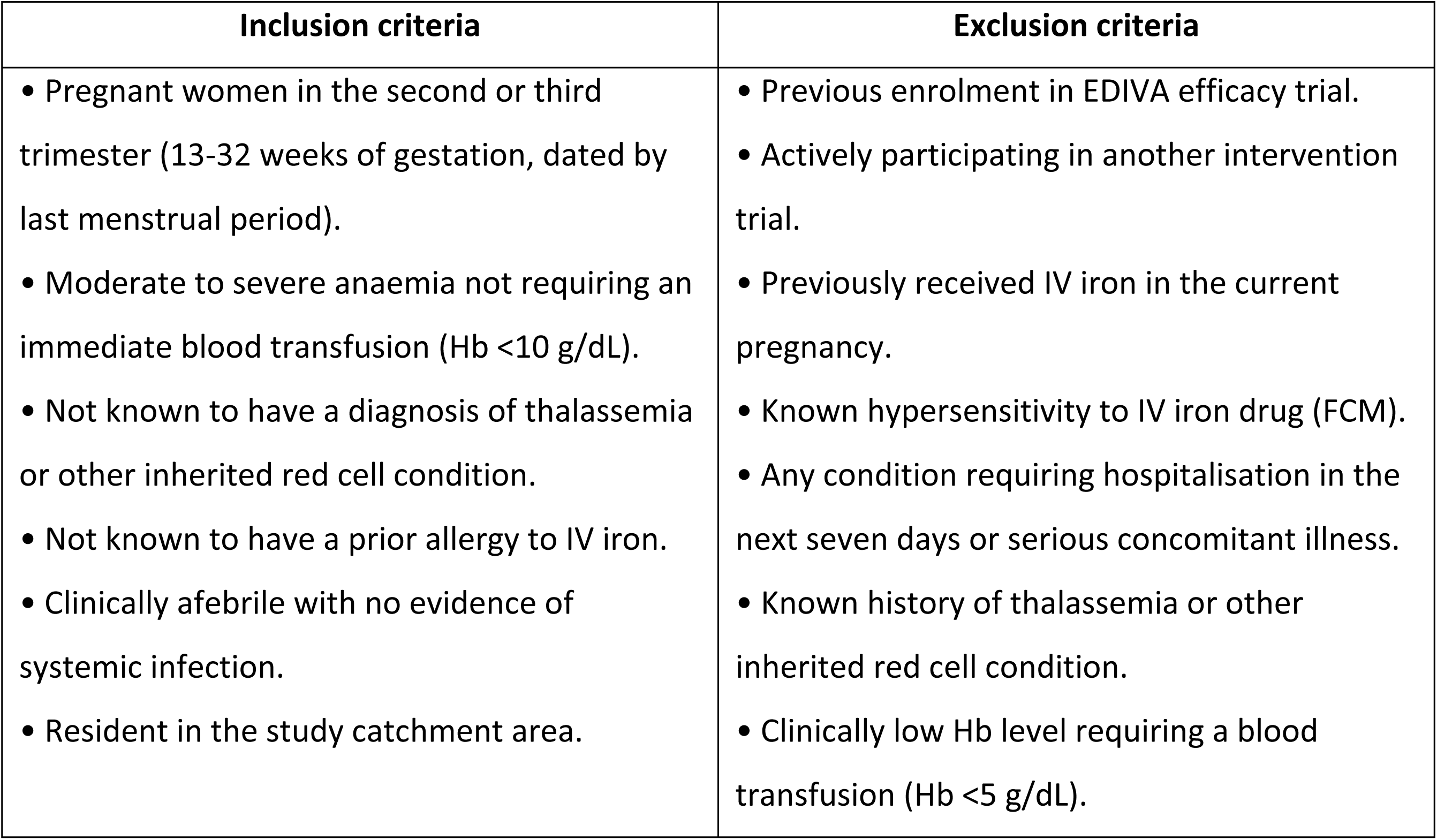
Inclusion and exclusion criteria for referral to IV iron.

| Inclusion criteria | Exclusion criteria |
| --- | --- |
| • Pregnant women in the second or third trimester (13-32 weeks of gestation, dated by last menstrual period).<br>• Moderate to severe anaemia not requiring an immediate blood transfusion (Hb <10 g/dL).<br>• Not known to have a diagnosis of thalassemia or other inherited red cell condition.<br>• Not known to have a prior allergy to IV iron.<br>• Clinically afebrile with no evidence of systemic infection.<br>• Resident in the study catchment area. | • Previous enrolment in EDIVA efficacy trial.<br>• Actively participating in another intervention trial.<br>• Previously received IV iron in the current pregnancy.<br>• Known hypersensitivity to IV iron drug (FCM).<br>• Any condition requiring hospitalisation in the next seven days or serious concomitant illness.<br>• Known history of thalassemia or other inherited red cell condition.<br>• Clinically low Hb level requiring a blood transfusion (Hb <5 g/dL). |

##### Implementation outcomes measures

###### Primary outcome

The primary feasibility outcome will be the proportion of pregnant women with identified moderate or severe anaemia in the second or third trimester that received IV iron in the demonstration project.

###### Secondary outcomes

The secondary feasibility outcomes will be adherence to key tasks across each care pathway including the percentage of eligible pregnant women in the second or third trimester screened for anaemia, diagnosed with moderate or severe anaemia and referred for IV iron treatment. Other secondary implementation outcomes are acceptability, fidelity, and costs. Data collection and analyses for the implementation outcomes are described in the phase 3 process evaluation section and in Table 2.

**Table 2.**
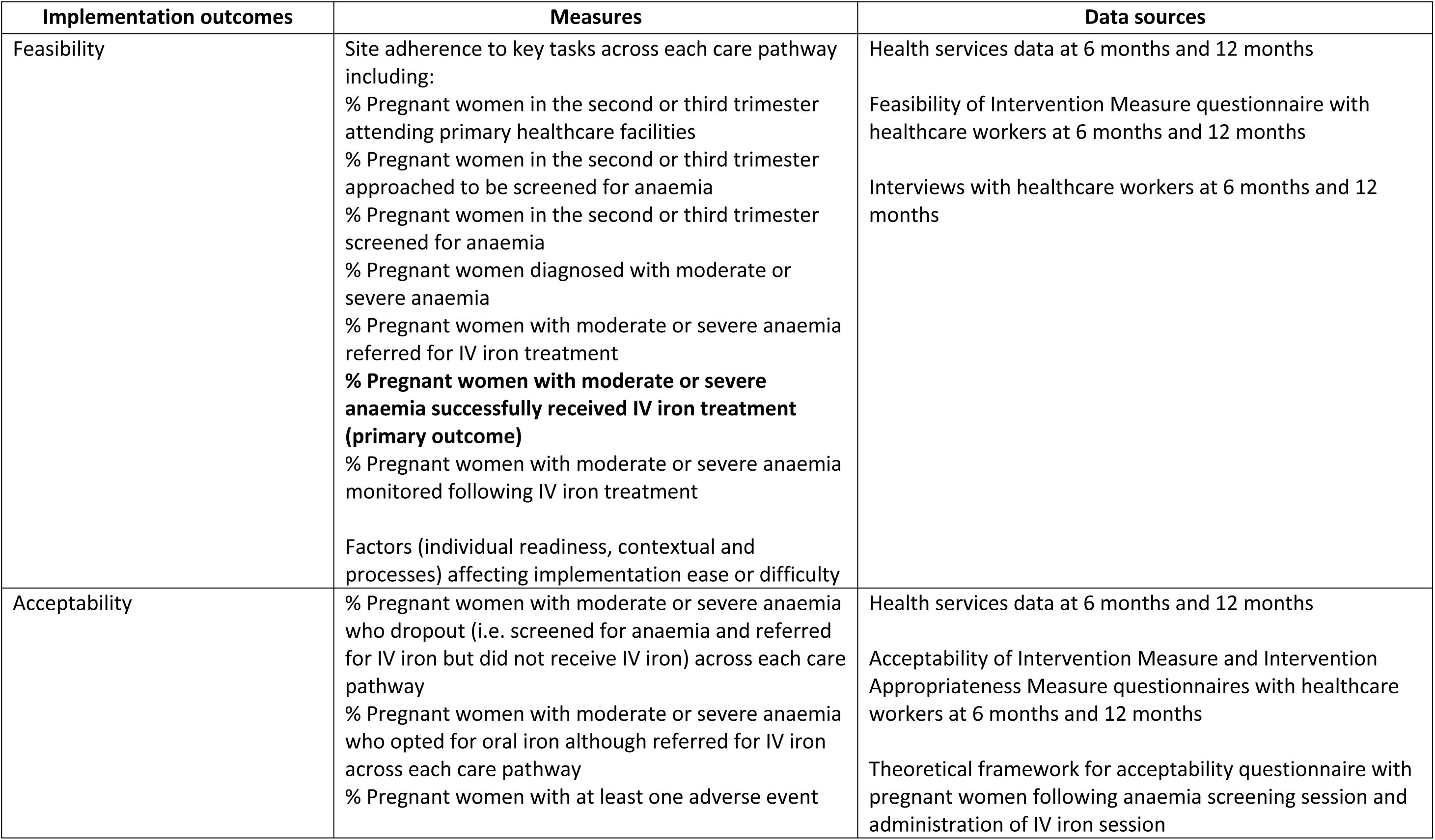
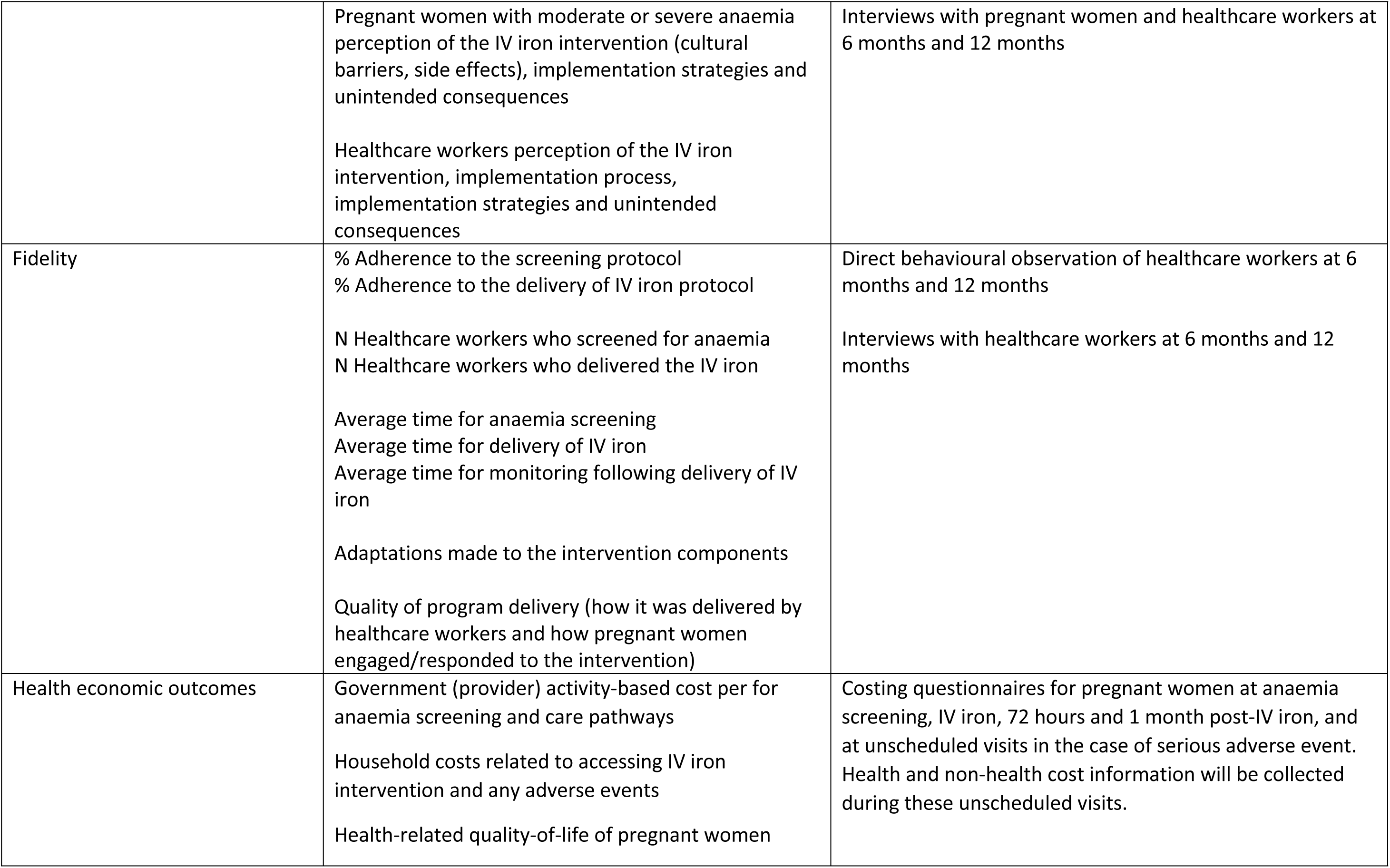
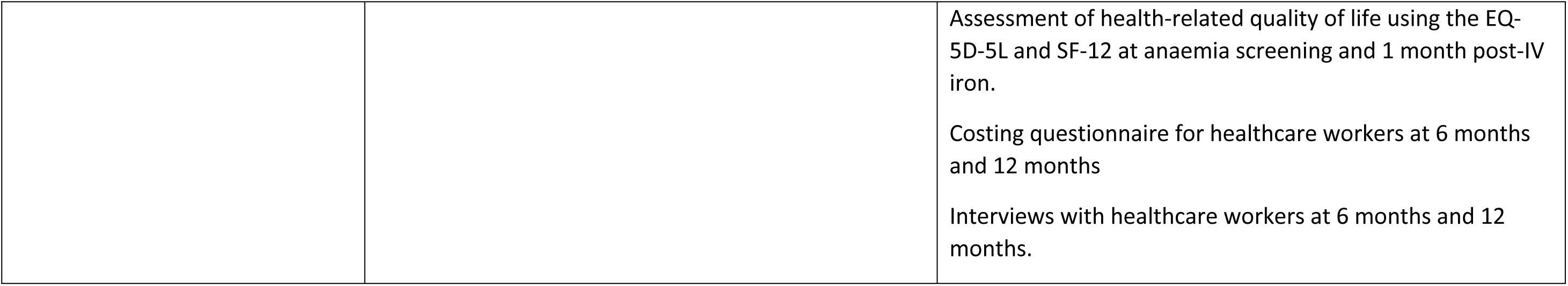
Implementation and health economic outcomes, measures and data sources for mixed methods data collection.

| Implementation outcomes | Measures | Data sources |
| --- | --- | --- |
| Feasibility | Site adherence to key tasks across each care pathway including:<br>% Pregnant women in the second or third trimester attending primary healthcare facilities<br>% Pregnant women in the second or third trimester approached to be screened for anaemia<br>% Pregnant women in the second or third trimester screened for anaemia<br>% Pregnant women diagnosed with moderate or severe anaemia<br>% Pregnant women with moderate or severe anaemia referred for IV iron treatment<br>**% Pregnant women with moderate or severe anaemia successfully received IV iron treatment (primary outcome)**<br>% Pregnant women with moderate or severe anaemia monitored following IV iron treatment<br>Factors (individual readiness, contextual and processes) affecting implementation ease or difficulty | Health services data at 6 months and 12 months<br>Feasibility of Intervention Measure questionnaire with healthcare workers at 6 months and 12 months<br>Interviews with healthcare workers at 6 months and 12 months |
| Acceptability | % Pregnant women with moderate or severe anaemia who dropout (i.e. screened for anaemia and referred for IV iron but did not receive IV iron) across each care pathway<br>% Pregnant women with moderate or severe anaemia who opted for oral iron although referred for IV iron across each care pathway<br>% Pregnant women with at least one adverse event | Health services data at 6 months and 12 months<br>Acceptability of Intervention Measure and Intervention Appropriateness Measure questionnaires with healthcare workers at 6 months and 12 months<br>Theoretical framework for acceptability questionnaire with pregnant women following anaemia screening session and administration of IV iron session |
|  | Pregnant women with moderate or severe anaemia perception of the IV iron intervention (cultural barriers, side effects), implementation strategies and unintended consequences<br>Healthcare workers perception of the IV iron intervention, implementation process, implementation strategies and unintended consequences | Interviews with pregnant women and healthcare workers at 6 months and 12 months |
| Fidelity | % Adherence to the screening protocol<br>% Adherence to the delivery of IV iron protocol<br>N Healthcare workers who screened for anaemia<br>N Healthcare workers who delivered the IV iron<br>Average time for anaemia screening<br>Average time for delivery of IV iron<br>Average time for monitoring following delivery of IV iron<br>Adaptations made to the intervention components<br>Quality of program delivery (how it was delivered by healthcare workers and how pregnant women engaged/responded to the intervention) | Direct behavioural observation of healthcare workers at 6 months and 12 months<br>Interviews with healthcare workers at 6 months and 12 months |
| Health economic outcomes | Government (provider) activity-based cost per for anaemia screening and care pathways<br>Household costs related to accessing IV iron intervention and any adverse events<br>Health-related quality-of-life of pregnant women | Costing questionnaires for pregnant women at anaemia screening, IV iron, 72 hours and 1 month post-IV iron, and at unscheduled visits in the case of serious adverse event. Health and non-health cost information will be collected during these unscheduled visits. |
|  |  | Assessment of health-related quality of life using the EQ-5D-5L and SF-12 at anaemia screening and 1 month post-IV iron.<br>Costing questionnaire for healthcare workers at 6 months and 12 months<br>Interviews with healthcare workers at 6 months and 12 months. |

###### Sample size

The sample size calculation for feasibility outcomes is based on two care pathways that will be implemented in the demonstration project over a one-year period, with an evaluation halfway to inform iteration of the pathways. The first month (transition period) is excluded from the calculation as the focus will be on introducing the intervention across the sites and address issues that may arise early on. We assume in a five-month implementation period that 470 and 356 pregnant women will be identified in the family welfare assistant’s register book for pathway 1 (e.g., North Bandar) and pathway 2 (e.g., South Bandar) respectively, and that 90% of these pregnant women will be screened for anaemia. Of these, we expect up to 13% of screened pregnant women will have moderate or severe anaemia (hb <10 g/dL) at one of these screenings (25) and thus approximately 55 and 42 women to be referred for IV iron infusion in each pathway, respectively. If an IV iron infusion is observed in 95% of all referrals (i.e., 52/55 and 40/42 respectively), then the 95% CI of the true underlying IV iron infusion rate among referrals is 85%-99% and 84%-99% respectively, using a binomial exact (Clopper-Pearson) method to obtain the two-sided 95% CI.

#### Implementation strategies

We anticipate at least one strategy for community members (including pregnant women) and two strategies for health workers (including comprehensive training for healthcare workers) will be implemented during the study period.

For community members, these strategies may aim to raise awareness of anaemia in pregnant women within the community, increase access to antenatal care services in government health facilities, and uptake of anaemia screening and treatment. Where possible, strategies will build on the existing strengths and opportunities within the community, such as religious leaders or existing community events.

For healthcare workers, one of the strategies will involve training to screen pregnant women for anaemia by measuring capillary blood using the HemoCue and to safely deliver IV iron in the primary healthcare setting. The anaemia screening training modules will focus on how to use and maintain the hemocues, aseptic procedures for the collection of capillary blood, identification of moderate and severe anaemia, and referral for IV iron administration. The IV iron administration training modules will focus on how to prepare the IV iron formulation dosage, how to perform IV cannulation using aseptic technique, safe management and disposal of waste, monitoring and responding to adverse and serious adverse events during IV iron infusion including referral to secondary care. Training will be delivered by the medical doctors in our research team and will be followed by competency-based evaluation. Refresher training will be conducted regularly. We will embed our research team in the primary healthcare system to oversee the administration of IV iron throughout the demonstration project and conduct monthly monitoring, but the intention will be that the healthcare workers lead all aspects of the intervention including anaemia screening of pregnant women, referral, and administration of IV iron.

### Phase three: Process evaluation

We will conduct a mixed methods process evaluation to explore the performance of the care delivery pathways guided by the MRC for developing and evaluating complex intervention framework (28, 29) and implementation outcomes measures as described by Proctor et al. (46). Our key implementation outcomes measures are feasibility, acceptability, and fidelity. Alongside the key implementation outcomes, we will assess the costs associated with the intervention implementation from a provider and patient perspective, through a micro-costing study. We will collect data on the health-related quality of life of pregnant women. These health economic outcomes will feed into a modelled economic evaluation comparing IV iron to oral supplementation using data from both the RCT and demonstration study. Together these implementation and health economic outcome measures can provide evidence about the expected financial feasibility of the intervention, and the likelihood that the intervention will be adopted into routine antenatal care practice.

#### Study participants and recruitment

Complete target population sampling will be used to identify and select eligible pregnant women and healthcare workers to participate in the process evaluation (47). All pregnant women participating in the demonstration project will be invited to complete questionnaires and sub-set of pregnant women, obtained through purposeful random sampling, will be invited to participate in an interview at 6 and 12-month post-implementation. The inclusion and exclusion criteria for participants will be reflective of the demonstration project criteria (Table 1). Similarly, all healthcare providers involved in the demonstration project at each site will be invited to complete questionnaires and participate in an interview at 6 months and 12 months post-implementation. Previous quantitative studies have shown less than 50% response rate for self-administered questionnaires (48). To ensure a high response rate and to address the low literacy level of pregnant women in Bandar (49) we will use interviewer-administered questionnaires where possible.

#### Data collection and analysis

Table 2 outlines the implementation outcomes measures, participant groups, and data collection to evaluate the performance of the care delivery pathways. We will apply a convergent mixed methods design (50) in which we will collect quantitative data to measure elements of feasibility and fidelity, and qualitative data to explore the acceptability of an IV iron intervention, how it worked, and why. We will conduct independent quantitative and qualitative data analysis. We will then triangulate the quantitative and qualitative findings to gain an in-depth understanding of feasibility and acceptability of an IV iron intervention by healthcare workers and pregnant women, and the contextual factors influencing the adoption and fidelity of delivery of such an intervention.

For feasibility and acceptability of an IV iron intervention, we will collect quantitative and qualitative data at two timepoints (6-months and 12-months post-implementation). For healthcare workers, feasibility and acceptability for anaemia screening and delivery of IV iron will be assessed using the Feasibility of Intervention Measure (FIM), Acceptability of Intervention Measure (AIM), and Intervention Appropriateness Measure (IAM) (51). For pregnant women, acceptability of being screened for anaemia and receiving IV iron will be assessed using the Theoretical Framework of Acceptability (TFA) questionnaire (52). To complement the quantitative findings, we will conduct semi-structured interviews with healthcare workers and pregnant women to explore their experiences and use of an IV iron intervention, factors affecting their perceived acceptability and feasibility including access barriers and enablers, and opportunities for improvement.

For fidelity of the intervention components, we will focus on adherence to anaemia screening and the administration of IV iron processes. Adherence will be assessed using direct behavioural observations of anaemia screening and the administration of the IV iron by members of our research team across all sites at 6-months and 12 months post-implementation using a procedural fidelity checklist. To complement the quantitative findings, we will conduct semi-structured interviews with healthcare workers to investigate the barriers and enablers to the fidelity of delivery of the intervention components. CFIR and TFA will be used to inform the development of all interview guides. All questionnaires and interview guides will be translated into Bengali.

Binomial exact (Clopper-Pearson) 95% Confidence Intervals (CIs) for a single proportion will be used to obtain CIs for the primary feasibility outcome (percentage of referrals that culminate in IV iron infusions) and each binary quantitative outcome; means and 95% CIs will be reported for secondary continuous quantitative outcomes (such as time for delivery of IV iron). Descriptive statistics will be used to summarise the characteristics of study participants. For qualitative data, we will apply the framework method using a mix of deductive and inductive thematic analysis to identify themes (35).

##### Ethical considerations

We obtained ethics approval for the demonstration project from the Research and Ethical Review Committee of icddr,b (PR-20125) and WEHI (21/5). We will seek informed written consent from all participants before participating in the implementation research activities and the demonstration project. The consent forms, questionnaires, and interview guides are available in Bengali and English.

##### Study timeline

The timeframe for the implementation research program is approximately 4 years. When this paper was submitted for publication, phase 1 formative research was completed. Phase 2 demonstration project is currently underway and participant recruitment commenced on the 16^th^ March 2024. Phase 3 process evaluation will be ongoing throughout the demonstration project, which is anticipated to be over 12 months. Report writing and result dissemination will occur following the completion of each phase.

## Discussion

In this paper, we describe the protocol for the formative research and a demonstration project that aims to demonstrate the real-world feasibility and acceptability of implementing an IV iron intervention to treat pregnant women with moderate and severe anaemia in the primary healthcare system of Bangladesh. Despite significant investments by the Bangladesh Ministry of Health and Family Welfare, including an oral IFA supplementation program since 2001, the reduction in the prevalence of anaemia in pregnant women has stagnated (15). Modern IV iron products are an alternative treatment available in HICs for women experiencing anaemia in pregnancy. Modern IV iron products, including FCM, have many benefits over oral IFA supplementation, and could potentially be implemented in the Bangladesh primary healthcare system. However, the acceptability and feasibility of the intervention will be contingent on several factors, including the culture of organisations and the socio-cultural beliefs of end-users (22, 53). In addition to the formative research on factors and strategies influencing the implementation of an IV iron program for pregnant women with anaemia, our research program includes a demonstration project. Through various care pathways, the demonstration project will test the implementation and scalability of an IV iron program for pregnant women with anaemia in the Bangladesh primary healthcare system.

### Expected study outcomes

The main outcome of this study is determining the real-world feasibility and acceptability of implementing an IV iron intervention to treat pregnant women with moderate and severe anaemia in the Bangladesh primary healthcare system. This will be achieved by implementing and evaluating various care pathways to support the uptake and delivery of an IV iron intervention in primary care. We will identify how many pregnant women with anaemia were screened and successfully received IV iron across each pathway, show how feasible and acceptable the IV iron intervention is, estimate the cost of implementing IV iron across the different anaemia screening and delivery pathways, and demonstrate to what extent the IV iron intervention was delivered as planned. The formative research will also provide a thorough understanding of 1) how anaemia in pregnant women is currently managed in Bangladesh, and potential health system barriers and enablers to implementing an IV iron intervention, and 2) culturally appropriate strategies to inform the implementation of an IV iron intervention in primary care. We anticipate that the demonstration project will increase attendance to antenatal care services for anaemia screening and that pregnant women with moderate and severe anaemia will have timely access to IV iron treatment. This will ultimately support a reduction in the prevalence of anaemia in pregnancy. Understanding the financial feasibility of implementing IV iron and can be used in the planned economic evaluation of IV iron compared with oral IFA for treatment of moderate or severe anaemia among pregnant women in rural Bangladesh. Findings from our research program if successful can be used by policymakers from the Ministry of Health and Family Welfare to inform sub-district, district, or nationwide scale-up of an IV iron intervention for pregnant women with anaemia.

### Anticipated problems and solutions

Government healthcare providers and management may be hesitant to support the implementation of an IV iron intervention for pregnant women experiencing anaemia in their workplace due to limited knowledge of the intervention and limited resources to deliver the intervention. We will provide educational information about the effectiveness and safety of FCM. The demonstration project is being preceded by a RCT comparing the effects of IV iron FCM to oral IFA supplementation on a range of maternal and child health outcomes in Rupganj and Sonargaon Upazilas (26), and the interim safety evidence from this RCT, and similar trials being conducted in other LMICs (54, 55, 56) will be presented. This will be further mitigated by extensively involving government healthcare providers in the formative research, with their opinions and preferences informing the co-design of the care pathways in the demonstration project. During the process evaluation, interviews will be conducted with government healthcare providers to identify and address any concerns they may have. There is also the potential that pregnant women may perceive IV iron negatively or hold misconceptions. Like government healthcare providers, pregnant women will be extensively involved in the co-design of the care pathways in the demonstration project, with findings from the formative research also informing community sensitisation activities and mobilisation.

### Applicability of the results

The results of Phase 1: Formative research (health system review, key stakeholder interviews, health facility readiness assessment) will inform the co-design workshops, which in turn will inform Phase 2: Demonstration project. Findings from Phase 1: Formative research, will ensure that the demonstration project is appropriate and applicable to the context of the Bangladesh primary healthcare system. Findings from Phase 2: Demonstration project, will inform understanding of which care pathway within the Bangladesh primary healthcare system is the most feasible and acceptable for implementing an IV iron intervention for pregnant women with moderate and severe anaemia, and the costs associated with the different care pathways. Our systematic approach to conducting the formative research and co-designing the demonstration project will increase the likelihood of the IV iron intervention being adopted, scaled and sustained across Bangladesh. Further, our findings may apply to other LMICs looking to implement an IV iron intervention for pregnant women experiencing anaemia.

### Dissemination of findings

Findings from the formative research and demonstration project will be regularly presented to the EDIVA Advisory Group, consisting of members from the Bangladesh Ministry of Health and Family Welfare and WHO. Findings will also be disseminated to key stakeholders through several outputs, as appropriate, including – evidence briefs, meetings with community members, journal articles, and relevant conferences.

## Data Availability

Protocol paper with no data.

## Abbreviations

AIM: Acceptability of Intervention Measure
CFIR: Consolidated Framework for Implementation Research
DGDA: Directorate General of Drug Administration
DGFP: Directorate General of Family Planning
DGHS: Directorate General of Health Services
EDIVA: Efficacy and Demonstration of IntraVenous Iron for Anaemia in pregnancy
FBC: Full blood count
FCM: Ferric carboxymaltose
FIM: Feasibility of Intervention Measure
HIC: High-income countries
Hb: hemoglobin
IAM: Intervention Appropriateness Measure
IFA: Iron and folic acid
IV: Intravenous
LMIC: Low- and middle-income countries
MRC: Medical Research Council
RCT: Randomised controlled trial
WHO: World Health Organization

## Declarations

### Ethics approval and consent to participate

Ethics approval for the demonstration project was obtained from the Research and Ethical Review Committee of icddr,b (PR-20125) and WEHI (21/5).

### Consent for publication

Not applicable

### Availability of data and materials

Not applicable

## Funding

This research program is supported by the Gates Foundation (INV-017316). The opinions expressed in this work are those of the authors alone and shall not be attributed to the Gates Foundation.

### Authors’ contributions

SP and JDM conceived the study. KHP and EV drafted the manuscript, with input from all authors. KHP, EV, HS and BKS developed the study tools. BKS and KHP led the formative research and will be leading the process evaluation with support from EV and HS. NC will lead the health economics evaluation with support from CG. All authors were involved in the selection of anaemia care pathways and implementation strategies. AM and SB calculated the sample size for the demonstration project. BKS and QR are responsible for the conduct of the demonstration project and implementation strategies, with support from SA, IH, ED, KHP, EV and HS. All authors read and approved the final version of the manuscript.

## Acknowledgement

We dedicate this manuscript to our beloved colleague, mentor and friend Professor Margaret Kelaher who played a key role in the design of the research program. She sadly passed away in March 2021.

